# Acute respiratory distress syndrome after SARS-CoV-2 infection on young adult population: international observational federated study based on electronic health records through the 4CE consortium

**DOI:** 10.1101/2022.03.31.22273257

**Authors:** Bertrand Moal, Arthur Orieux, Thomas Ferté, Antoine Neuraz, Gabriel A Brat, Paul Avillach, Clara-Lea Bonzel, Tianxi Cai, Kelly Cho, Sébastien Cossin, Romain Griffier, David A Hanauer, Christian Haverkamp, Yuk-Lam Ho, Chuan Hong, Meghan R Hutch, Jeffrey G Klann, Trang T Le, Ne Hooi Will Loh, Yuan Luo, Adeline Makoudjou, Michele Morris, Danielle L Mowery, Karen L Olson, Lav P Patel, Malarkodi J Samayamuthu, Fernando J Sanz Vidorreta, Emily R Schriver, Petra Schubert, Guillaume Verdy, Shyam Visweswaran, Xuan Wang, Griffin M Weber, Zongqi Xia, William Yuan, Harrison G Zhang, Daniela Zöller, Isaac S Kohane, The Consortium for Clinical Characterization of COVID-19 by EHR (4CE), Alexandre Boyer, Vianney Jouhet

## Abstract

**Purpose:** In young adults (18 to 49 years old), investigation of the acute respiratory distress syndrome (ARDS) after severe acute respiratory syndrome coronavirus 2 (SARS-CoV-2) infection has been limited. We evaluated the risk factors and outcomes of ARDS following infection with SARS-CoV-2 in a young adult population.

**Methods:** A retrospective cohort study was conducted between January 1st, 2020 and February 28th, 2021 using patient-level electronic health records (EHR), across 241 United States hospitals and 43 European hospitals participating in the Consortium for Clinical Characterization of COVID-19 by EHR (4CE). To identify the risk factors associated with ARDS, we compared young patients with and without ARDS through a federated analysis. We further compared the outcomes between young and old patients with ARDS.

**Results:** Among the 75,377 hospitalized patients with positive SARS-CoV-2 PCR, 1001 young adults presented with ARDS ( 7.8% of young hospitalized adults). Their mortality rate at 90 days was 16.2% and they presented with a similar complication rate for infection than older adults with ARDS. Peptic ulcer disease, paralysis, obesity, congestive heart failure, valvular disease, diabetes, chronic pulmonary disease and liver disease were associated with a higher risk of ARDS. We described a high prevalence of obesity (53%), hypertension (38%-although not significantly associated with ARDS), and diabetes (32%).

**Conclusion:** Trough an innovative method, a large international cohort study of young adults developing ARDS after SARS-CoV-2 infection has been gather. It demonstrated the poor outcomes of this population and associated risk factor.

## 1 Introduction

Acute respiratory distress syndrome (ARDS)[1], is a frequent complication after severe acute respiratory syndrome coronavirus 2 (SARS-CoV-2) infection. According to studies, it appears in 3.4% of the population with a laboratory positive PCR confirmation of infection to the SARS-CoV-2[2], up to 31% of hospitalized patients[3–5], and 92% of patients admitted to the intensive care unit[4](ICU).

ARDS has a severe impact on patient outcomes. In a cohort study carried out in New York City on COVID-19 patients, the mortality of ARDS patients reached 39%[4]. ARDS has been frequently associated with long-term disabilities[6–10] and represents a heavy care burden for health systems[11] due to long ICU stays and extended rehabilitation[7,9].

Age is an important risk factor for developing ARDS[3]. However, young adults (18-49 years old) represented a third of hospitalized patients[12] and a quarter of patients admitted to the ICU[4]. Based on the Premier Healthcare Database, which includes 1,030 hospitals in the United States, Cunningham et al.[3] reported that 21% of young adults (aged 18 to 34 years) hospitalized with COVID-19 disease were admitted to the ICU and 10% required mechanical ventilation. Similarly, in a separate cohort, young adults represented more than 20% of the patients admitted to ICUs for COVID-19 infection with ARDS[3].

Few studies[12–15] have investigated the young adult population, mostly were single-center analyses, all exclusively in the U.S. population and none focused on ARDS patients. To our knowledge, there have been no specific studies on ARDS after SARS-CoV-2 infection in the young adult population among an international cohort. This may be due to the difficulty in obtaining a large sample of this population. Key questions remain related to the risk factors of ARDS in young adults, and the difference, in terms of outcomes, compared to an older population. In this study, we investigate the risk of ARDS among young adults hospitalized with COVID-19 using an international cohort from the international Consortium for Clinical Characterization of COVID-19 <u>(4CE)</u>[16–21]. This international consortium collects data from 342 hospitals in 6 countries and develops an innovative federated approach for electronic health records (EHR) analysis.

Through a federated analysis, the objectives were to evaluate the risk factors for developing ARDS following infection with SARS-CoV-2 and hospitalization in young adults and to compare characteristics, care, and outcomes between this population and an older population (greater than 49 years old) who similarly developed ARDS during their COVID-19 hospitalization.

## 2 Patients and Methods

The 4CE consortium [16–21] has developed a framework to extract and standardize data directly from the EHRs of participating healthcare systems (HS) and to streamline federated analyses without sharing patient-level data. A common data model for structuring patient-level data was adopted to enable identical analyses across all participating HS. Figure 1 presents the workflow from 4CE data collection to ARDS analysis.

**Figure 1.**
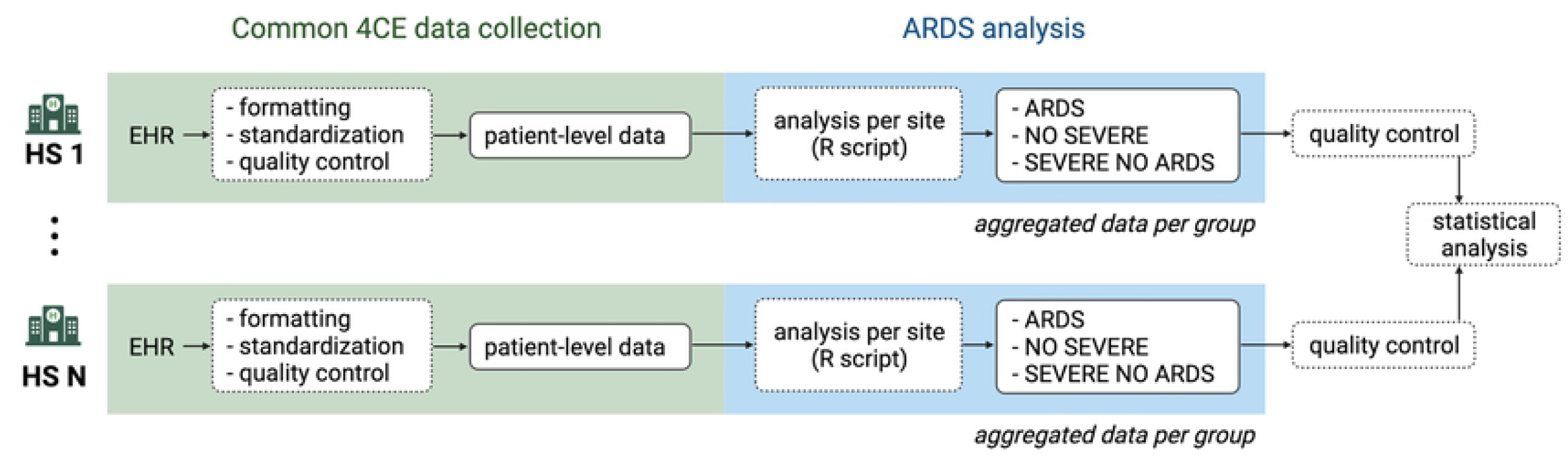
Study workflow. From EHR extraction to ARDS analysis on aggregated data (HS: healthcare system).

### 2.1 Common 4CE Data collection by HS

As previously described[16], each participating HS were responsible for and obtained ethics approval, as needed, from the appropriate ethics committee at their institution. IRB protocols were reviewed and approved at APHP (IRB00011591, Project CSE-20-29_ClinicalCOVID), Bordeaux University Hospital (Registration #CHUBX2020RE0253), Mass General Brigham (IRB#2020P001483), Northwestern University (IRB# STU00212845), University of Kansas (STUDY00146505), University of Freiburg (Application #255/20, Process #210587), and at VA North Atlantic, Southwest, Midwest, Continental, and Pacific (IRB # 3310-x).

The research was determined to be exempt at University of Michigan (IRB# HUM00184357), Beth Israel Deaconess Medical Center (IRB# 2020P000565), University of Pittsburgh (STUDY20070095), and University of Pennsylvania (IRB#842813). University of California Los Angeles determined that this study does not need IRB approval because research using limited data sets does not constitute human subjects research.

#### 2.1.1 Cohort identification

Across each participating HS, we included all hospitalized patients within 7 days before and up to 14 days after a positive PCR SARS-CoV-2 test. The first hospital admission date within this time window was considered day 0 (the index date). Note that although all patients had a positive PCR test near their admission date, it is possible that for some patients the hospitalization was for reasons other than COVID-19.

#### 2.1.2 Patient-level data collection by HS

Patient-level data were collected by HSs, which can represent one or several hospitals. At each HS, data were extracted directly from the EHR and consisted of time to admission and discharge, survival status, sex and age group [(18−25, 26−49, 50−69, 70−79, and 80+ years old]. Diagnoses were collected from the first 3 digits of the billing code using international classification disease (ICD) version 10. This 3-digit rollup was adopted to account for finer-grained differences in coding practices across hospitals. Procedures related to endotracheal tube insertion or invasive mechanical ventilation were collected and were denoted as severe procedures[17]. Medications administered were collected at the class level (as per the ATC standard nomenclature[22], e-Appendix 1). Severe medication[17] refers to sedatives/anesthetics or treatment for shock (classes: SIANES, SICARDIAC).

All patient-level data were standardized to a common format, then stored and analyzed locally at each HS. Several quality controls were conducted iteratively at each HS to ensure the quality of the data.

### 2.2 ARDS analysis

#### 2.2.1 Data aggregation by HS for ARDS analysis

Final data extraction was completed on 30^th^ August 2021 and included patient hospitalizations occurring from 1^st^ January 2020 to 28^th^ February 2021. All patients of 18 years or older were included in the analysis. ARDS patients were identified using the ICD10 code, J80 - Acute respiratory distress syndrome.

Using patient-level data, each HS ran an R script locally to classify patients into 3 groups as follows:

- ARDS: Patients with an ARDS ICD code
- NO_SEVERE: Patients without an ARDS ICD code, severe medication or severe procedure
- SEVERE_NO_ARDS: Patients with severe medication or severe procedure but without an ARDS ICD code

For the analysis, the cohort was divided into two age groups: patients aged 18 to 49 years and patients older than 49 years (Figure 2). For each group, the number of patients was aggregated in terms of:

**Figure 2.**
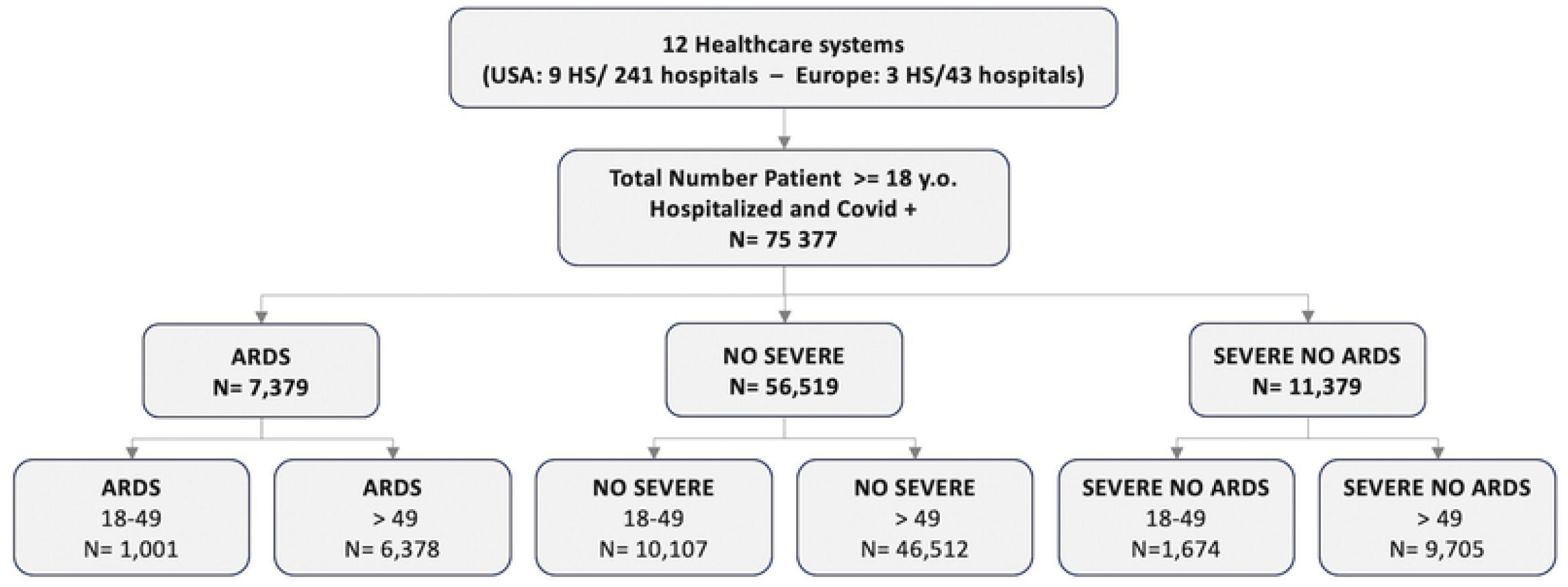
: Flow chart, distribution of patients per group (y.o.= years old)

- Age, sex, mortality at 90 days after the admission
- Each ICD code, Elixhauser index(23) and complication class (e-Appendix 2-3)

Aggregate data were centrally collected, and several quality controls were executed before pooling the aggregated data together. Descriptive analysis was presented e-Table 1.

#### 2.2.2 Statistical Analysis

##### 2.2.2.1 Risk factor: comparison between young patients with and without ARDS

To identify the risk factors associated with an ARDS after SARS-CoV-2 infection and hospitalization, we compared the young patients with ARDS and the young non severe patients. Patients classified in the “SEVERE_NO_ARDS” group were excluded from this analysis. For comorbidities classified by the Elixhauser Comorbidity Index[23], risk ratios with confidence intervals were calculated from a univariable analysis considering diagnoses recorded between 365 days before (−365) the admission and 90 days after (+90) the admission. First univariable analysis was performed at each HS and aggregated through a random effect meta-analyses to account for heterogeneity between HS. In addition, comorbidities associated with ARDS in this meta univariable analysis and sex were selected for a multivariable analysis. Multivariable analysis was performed at each HS and then aggregated through another meta-analysis with random effect.

##### 2.2.2.2 Complications and mortality: comparison between young and old adults with ARDS

The proportion of patients per sex were evaluated and compared between young adults and older adults with ARDS. Complications were identified as novel diagnoses established between the day of admission and +90 days after the admission. To compare complications between young and older patients with ARDS, we performed a univariable analysis and reported estimated risk ratios with confidence intervals. Moreover, mortality was evaluated for both groups at 90 days after the index admission.

Statistical analyses were performed locally at each HS and then aggregated via meta-analysis with the R package metafor[24].

## 3 Results

12 HS participated in the analysis: 9 U.S. HS representing 241 hospitals, two French HS representing 42 hospitals, and one German HS representing 1 hospital (Table 1). 75,377 hospitalized patients with biological confirmation of COVID were included in the analysis. About 7.8% (1001/12,782, HS range: 1.6 to 15%) of hospitalized young adults with COVID developed ARDS compared to 10.2% (6378/62595, HS range: 1.8 to 21.2%) of older patients. Young patients represented 13.4% (1001/7379) of ARDS patients (HS range: 6.5% to 24.5%).

**Table 1:** Name, City, Country, Number of hospitals per HS, number of Beds and Inpatient discharges/year per HS

| Healthcare System | City | Country | Hospitals | Beds | Inpatient discharges/year |
| --- | --- | --- | --- | --- | --- |
| Assistance Publique - Hôpitaux de Paris | Paris | France | 39 | 20,098 | 1,375,538 |
| Bordeaux University Hospital | Bordeaux | France | 3 | 2,676 | 130,033 |
| Medical Center, University of Freiburg | Freiburg | Germany | 1 | 1,66 | 71,5 |
| Beth Israel Deaconess Medical Center | Boston, MA | USA | 1 | 673 | 40,752 |
| Mass General Brigham (Partners Healthcare) | Boston, MA | USA | 10 | 3,418 | 163,521 |
| University of Pennsylvania | Philadelphia, PA | USA | 5 | 2,469 | 118,188 |
| University of Michigan | Ann Arbor, MI | USA | 3 | 1 | 49,008 |
| Northwestern University | Chicago, IL | USA | 10 | 2,234 | 103,279 |
| University of California, LA | Los Angeles, CA | USA | 2 | 786 | 40,526 |
| University of Pittsburgh / UPMC | Pittsburgh, PA | USA | 39 | 8,085 | 369,3 |
| University of Kansas Medical Center | Kansas City, KS | USA | 1 | 794 | 54,659 |
| Veteran affairs | Multiple cities | USA | 170 | 13,801 | 680,687 |

### 3.1 Risk factors: comparison between young adults with ARDS and young non severe patients (Table 2)

For the risk factor analysis, young ARDS patients (n = 1001) were compared to young non severe patients (n = 10,107). Among young ARDS patients, 43/1001 (4.3%) were aged between 18 to 25 years old. In an univariable analysis, patients aged 26 to 49 years old had an increased risk of developing ARDS compared to those aged 18 to 25 years old (RR = 2.94; 95% CI: [2.11; 4.1]). Due to the low proportion of patients between 18 to 25 years, age class was not included in the multivariable analysis.

**Table 2:** Number and percentage of patients per age groups, per sex, per Elixhauser comorbidities for young adult patients with ARDS and non severe young adult patients. Risk ratio associated in uni- and multivariable analysis

| Variables | ARDS | NO SEVERE | Univariable analysis |  | Multivariable analysis |  |
| --- | --- | --- | --- | --- | --- | --- |
|  | ages 18-49<br>n = 1001<br>n (%) | ages 18-49<br>n = 10107<br>n (%) | Risk Ratio with CI<br>(95%) | p-value | Risk Ratio with CI<br>(95%) | p-value |
| Age groups, reference: 18 to 25 years old |  |  |  |  |  |  |
| 18to25 | 43 (4.3) | 1207 (11.9) | 2.9 [ 2.1; 4.1] | <0.001 | not include |  |
| 26to49 | 966 (96.5) | 8900 (88.1) |  |  |  |  |
| Sex, reference: female |  |  |  |  |  |  |
| female | 327 (32.7) | 4427 (43.8) | 1.7 [1.3; 2.2] | <0.001 | 1.71 [1.2; 2.4] | 0.003 |
| male | 672 (67.1) | 5680 (56.2) |  |  |  |  |
| Comorbidities (Elix Hauser class), ICD code from -365 days before to + 90 days after admission |  |  |  |  |  |  |
| AIDS/HIV | 12 (1.2) | 121 (1.2) | 1 [0.5; 1.9] | 0.987 | not include |  |
| Alcohol abuse | 59 (5.9) | 895 (8.9) | 1 [0.7; 1.4] | 0.92 | not include |  |
| Cancer | 37 (3.7) | 280 (2.8) | 1.3 [0.9; 1.7] | 0.164 | not include |  |
| Chronic pulmonary disease | 219 (21.9) | 1406 (13.9) | 1.8 [1.6; 2.1] | <0.001 | 1.6 [1.3; 2.0] | <0.001 |
| Congestive heart failure | 143 (14.3) | 532 (5.3) | 3.4 [2.6; 4.4] | <0.001 | 2.2 [1.4; 3.6] | 0.001 |
| Diabetes | 322 (32.2) | 1691 (16.7) | 2.5 [2; 3.1] | <0.001 | 1.9 [1.4; 2.4] | <0.001 |
| Drug abuse | 65 (6.5) | 828 (8.2) | 1 [0.8; 1.3] | 0.997 | not include |  |
| Hypertension | 382 (38.2) | 2274 (22.5) | 2.5 [2; 3.2] | <0.001 | 1.4 [0.98; 1.9] | 0.062 |
| Hypothyroidism | 45 (4.5) | 431 (4.3) | 1.4 [1; 2.1] | 0.077 | not include |  |
| Liver disease | 179 (17.9) | 960 (9.5) | 2.1 [1.6; 2.8] | <0.001 | 1.6 [1.1; 2.3] | 0.01 |
| Obesity | 533 (53.2) | 2759 (27.3) | 2.9 [2.2; 3.9] | <0.001 | 2.8 [2.0; 4.0] | <0.001 |
| Paralysis | 64 (6.4) | 162 (1.6) | 2.9 [2.3; 3.6] | <0.001 | 3.7 [2.5; 5.5] | <0.001 |
| Peptic ulcer disease | 36 (3.6) | 60 (0.6) | 4.2 [2.9; 6] | <0.001 | 3.7 [2.1; 6.5] | <0.001 |
| Peripheral vascular disease | 37 (3.7) | 184 (1.8) | 2.7 [1.7; 4.2] | <0.001 | 1.2 [0.7; 2.1] | 0.485 |
| Psychoses | 52 (5.2) | 599 (5.9) | 1.1 [0.8; 1.4] | 0.513 | not include |  |
| Renal failure | 131 (13.1) | 590 (5.8) | 2.4 [1.9; 2.9] | <0.001 | 1.3 [0.9; 1.8] | 0.158 |
| Valvular disease | 92 (9.2) | 346 (3.4) | 2.9 [2.1; 4] | <0.001 | 1.9 [1.1; 3.3] | 0.025 |

In the multivariable analysis, compared to women, men had a higher risk for developing ARDS (RR = 1.71; 95% CI: [1.20; 2.43]) and the following comorbidities were significantly associated with ARDS: Peptic ulcer disease (RR = 3.66; 95% CI: [2.01; 6.49]), Paralysis (RR = 3.73; 95% CI:[2.52; 5.51]), Obesity (RR = 2.82; 95% CI: [2.06; 3.95]), Congestive heart failure (RR = 2.2; 95% CI: [1.36; 3.57]), Valvular disease (RR = 1.89; 95% CI: [1.08; 3.29]), Diabetes (RR = 1.85; 95% CI: [1.44; 2.38]), Chronic pulmonary disease (RR = 1.62; 95% CI: [1.34; 1.96]) and Liver disease (RR = 1.61; 95% CI: [1.12; 2.31]). Hypertension was not significantly associated (RR = 1.36 [0.98; 1.89]).

Peripheral vascular disease, and renal failure were associated with developing ARDS in univariable analysis, but not in multivariable analysis. AIDS/HIV, alcohol abuse, cancer, drug abuse, hypothyroidism, and psychosis were not associated with higher risk. Nicotine dependency was not associated with a higher risk (p= 0.138).

In the young ARDS population, we observed a high prevalence of comorbidities including obesity 533/1001 (53.3%), diabetes 382/1001 (38.2%), and hypertension 322/1001 (32.2%).

### 3.2 Complications and mortality: comparison between young and old adult population with ARDS

6378 patients aged > 49 with ARDS were compared to the young adult population with ARDS. The percentage of males was 67.1% (672/1001) and 75.2% (4797/6378) for the young population and the old population, respectively, without significant difference (p= 0.457).

#### 3.2.1 Complications(Table 3)

Young ARDS patients had a lower risk of developing the following complications: Acute kidney failure (RR = 0.76; 95% CI: [0.68; 0.85]); cardiac rhythm/conduction disorder (RR = 0.59; 95% CI: [0.47; 0.73]), disorders of fluid, electrolyte and acid-base balance (RR = 0.95; 95% CI: [0.88; 0.99]); and stroke (RR = 0.35; 95% CI: [0.23; 0.53]). However, they had a higher risk of developing pneumonia due to Streptococcus pneumoniae (RR = 1.78; 95% CI:1[1.16; 2.75]), and Streptococcal sepsis (RR = 1.58; 95% CI: [1.08; 2.31]). More than half of the young ARDS patients had Respiratory bacterial superinfection (538/1001 (53.8%)) during their hospitalization. No significant differences were found for the occurrence of pulmonary embolism (p = 0.671), affecting one in 10 patients in both groups with ARDS.

**Table 3:**
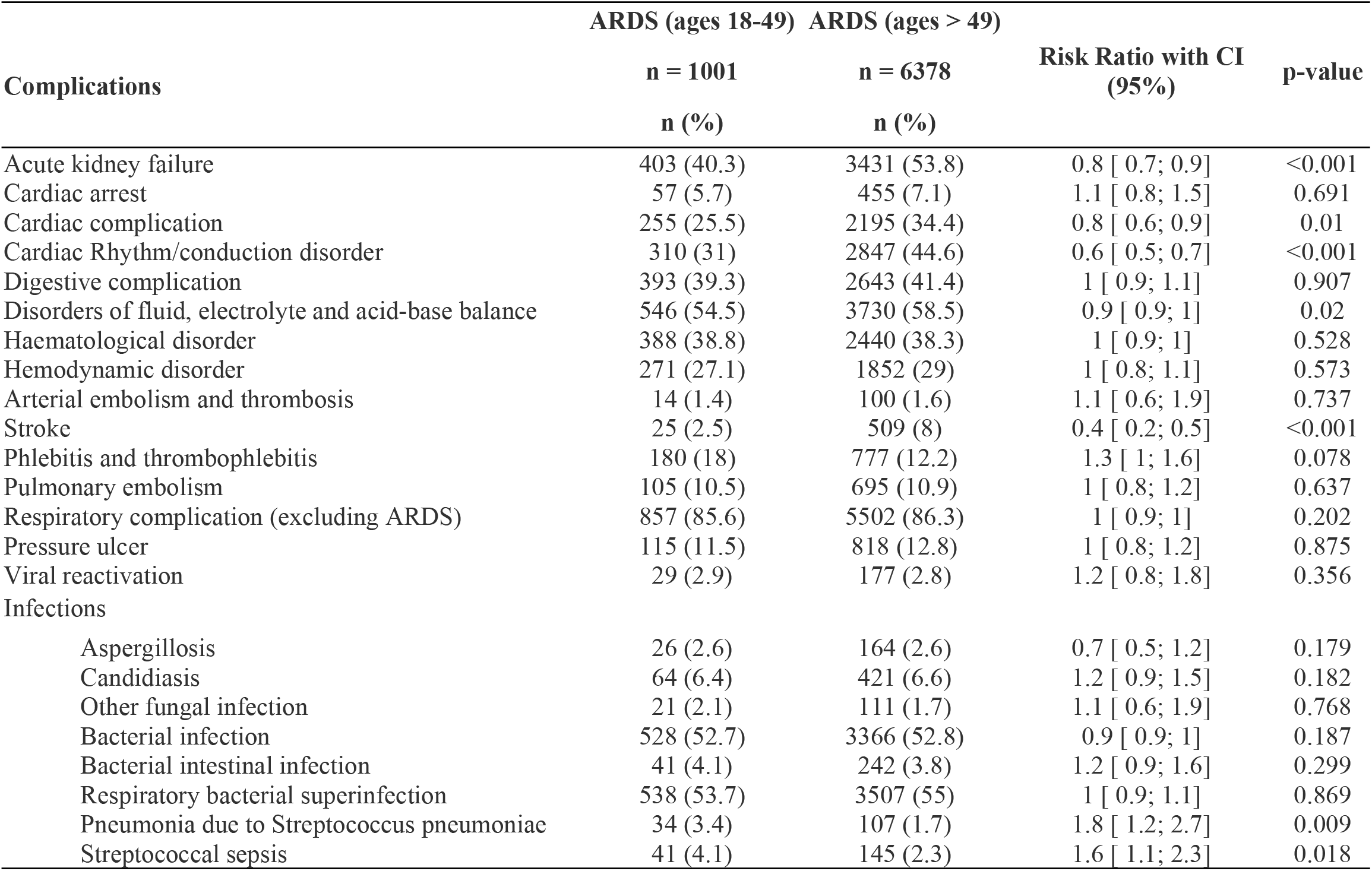
Proportion and associated risk ratio of complication classes for the young compared to old adult with ARDS

### 3.3 Mortality

90 days after admission, 16.2% (162/1001) of the young ARDS patients were deceased (HS range [11.2%; 36.8%]). In the older adult population with ARDS patients, the mortality was 41.1% (2619/6378, HS range [24.3%; 76.7%]).

## 4 Discussion

In a large international EHR-based cohort, we employed a novel federated approach including 241 hospitals in the United States and 43 in Europe, to describe comorbidities, complications, and mortality of young adults developing ARDS after SARS-CoV-2 infection. Even though young patients with ARDS represent a small proportion of hospitalized patients with COVID (HS range: [0.4%; 3.3%]), we were able to gather a large cohort thanks to this innovative method and demonstrated the poor outcome of young ARDS patients with notable mortality (16.2%).

### 4.1 Mortality and Complications

Independently on the etiology, in-hospital mortality for ARDS patients has been reported to be between 30 to 40%[7,25,26]. Mortality at 30 days for ARDS patients of any age with COVID-19 was reported at 39%[4] and corresponds to the mortality for the older ARDS population in our study. The young ARDS population’s mortality at 90 days was smaller, around 16.2% with large variability between HS [11.2; 36.8%], but it appeared high for this young population. In a 2018 study conducted in France, all-cause mortality of ICU patients in the same age range was estimated to be less than 10%[27]. The relatively higher risk of developing pneumonia due to Streptococcus pneumoniae and Streptococcal sepsis in young adults is probably related to their greater survival rate compared to older patients. The high frequency of complications in this young population emphasizes the major impact of ARDS on poor outcomes and mortality.

### 4.2 Risk factors

Although the proportion of the general population is low, ARDS appears in 7.8% of young hospitalized adults with COVID. These percentages are in agreement with those reported by Cummings et al.[3] and Cummingham et al.[13]. Among those young ARDS patients only 4,3% were aged between 18- and 25-years old. Patients developing ARDS in this young adult population had a high prevalence of obesity (53%), hypertension (38%) and diabetes (32%).

A limitation of relying on billing codes to identify comorbidities is the challenge of accurately distinguishing comorbidities from complications. In our analysis, comorbidities were considered as those diagnoses from billing codes assigned up to one year before and up to 90 days after the admission. This approach is more sensitive, but it can lead to considering complications as comorbidities. It is particularly true for peptic ulcer disease or paralysis which was identified as a comorbidity associated with ARDS but which is also known to be a common complication of mechanical ventilation[28,29] or prolonged ICU admission. We perform a complementary univariable analysis on the sub population who had previous hospital visits and considering only the ICD code related to those previous visits as comorbidities (one year and – 14 days before the admission). In this univariable analysis presented in e-Table 2, ARDS was associated with the presence of peptic ulcer disease or paralysis in a previous hospitalization, which explained our choice to keep both in the main multivariable analysis that means considering them as comorbidities. “Paralysis” regroups is related a large diversity of diagnoses. including encephalitis, myelitis and encephalomyelitis, hereditary ataxia, cerebral palsy, hemiplegia and hemiparesis, paraplegia (paraparesis) and quadriplegia (quadriparesis), and other paralytic syndromes (e-Appendix 2); but a common co-occurrence is reduced lung capacity which could contribute to its association with ARDS. The association with peptic ulcer as comorbidities remains unclear and requires additional investigations.

Obesity has been identified as a risk factor for poor outcome for ARDS[30] and for SARS-CoV-2 infection[3,14,15,31] and it also appears in this analysis as a risk factor in this young adult population. Diabetes has a controversial association with ARDS[32–34] but appears in this population as a risk factor and has also been associated with the severity of SARS-CoV-2 infection in other studies [3,14,15,31]. Despite its association with poor outcomes in several cohorts of COVID-19 patients[2,15], hypertension was not significantly associated with ARDS in our study, possibly due to the choice of the variable included in the multivariable analysis and/or a lack of power.

Congestive heart failure, valvular disease, chronic liver disease, and chronic pulmonary disease are not associated with ARDS in the literature, however, their associations with COVID-19 have been identified as a risk factor for poor outcomes [3,14,15,31]. Through our analysis, it seems that most of the comorbidities associated with ARDS in the young adult population are similar to the ones associated with poor outcomes after SARS-CoV-2 infection in the general population. However, for most of them, it is unclear whether they are truly related to the onset of ARDS or just general comorbidities. Further analysis needs to be carried out to eliminate confounding factors and better understand the potential mechanisms of those associations.

### 4.3 Limitations

Our major limitation is that group membership, comorbidities, and complication analyses are based on billing codes, procedures, and medications directly extracted from EHR. Variation in billing coding practices, especially across international healthcare systems, may result in missing data and related biases[35]. However, multiple quality controls have been established to reduce those potential biases. For the detection of ARDS patients, a correct sensibility is expected as billing code is related to reimbursement in most countries and ARDS is associated with heavy care.

To identify comorbidities associated with ARDS following hospitalization with COVID, a comparison was performed considering only non severe patients. Patients with mechanical ventilation, sedatives/anesthetics, or treatment for shock but without ARDS code were not included, which could generate a selection bias. This choice was conducted to eliminate potential miscoded ARDS patients and patients with severe disease or care not related to SARS-CoV-2 infection but with a concomitant infection. In addition, we believe that the descriptive analysis of the SEVERE_NO_ARDS brings credit to this choice (e-Table 1). Compared to the other groups, SEVERE_NO_ARDS population had the higher percentage of women (52.2%) and of patients with previous contact with the healthcare system (72%). In addition, 15.1% of those patients had a billing code associated with pregnancy and 36.1% with long-term drug therapy. These results suggest that the COVID-19 infection was simply concomitant but not the main cause of these hospitalizations.

## Conclusion

We federated a large EHR-based international cohort of young adults developing ARDS after COVID-19. ARDS appears in 7.8% of hospitalized young patients with COVID and was associated with high mortality (16.2%). Young adults developing ARDS presented a high prevalence of comorbidities, particularly obesity, hypertension (although not being associated with ARDS), and diabetes. ARDS development was associated with peptic ulcer disease, paralysis, obesity, congestive heart failure, valvular disease, diabetes, chronic pulmonary disease, and liver disease.

## Data Availability

Aggregated data presented in the study are available from XXXXX

## Glossary

SARS-CoV-2: severe acute respiratory syndrome coronavirus 2
ARDS: acute respiratory distress syndrome
EHR: electronic health records
4CE: Consortium for Clinical Characterization of COVID-19 by HER
ICU: intensive care unit
HS: healthcare systems
ICD: international classification diseases

